# BMI is positively associated with accelerated epigenetic aging in twin pairs discordant for BMI

**DOI:** 10.1101/2021.03.11.21253271

**Authors:** Sara Lundgren, Sara Kuitunen, Kirsi H. Pietiläinen, Mikko Hurme, Mika Kähönen, Satu Männistö, Markus Perola, Terho Lehtimäki, Olli Raitakari, Jaakko Kaprio, Miina Ollikainen

## Abstract

**Background:** Obesity is a heritable complex phenotype which can increase the risk of age-related outcomes. Biological age can be estimated from DNA methylation (DNAm) using various “epigenetic clocks.” Previous work suggests individuals with elevated weight also display accelerated aging, but results vary by epigenetic clock and population. Here, we utilize the new epigenetic clock GrimAge, which closely relates with mortality.

**Objectives:** We aimed to assess the cross-sectional association of BMI with age acceleration in twins to limit confounding by genetics and shared environment.

**Methods and Results:** Participants were from the Finnish Twin Cohort (FTC; n = 1424), including monozygotic (MZ) and dizygotic (DZ) twins, and DNAm was measured using the Illumina 450k array. Multivariate linear mixed effects models including MZ and DZ twins showed an accelerated epigenetic age of 1.02 months (*p*-value = 6.1 × 10^−12^) per 1-unit BMI increase. Additionally, heavier twins in a BMI-discordant MZ twin pair (ΔBMI > 3 kg/m^2^) had an epigenetic age 5.2 months older than their lighter co-twin (*p*-value = 0.0074). We also found a positive association between log(HOMA-IR) and age acceleration, confirmed by a meta-analysis of the FTC and two other Finnish cohorts (overall effect = 0.45 years, *p*-value = 0.0025) from adjusted models.

**Conclusion:** We identified significant associations of BMI and insulin resistance with age acceleration based on GrimAge, which were not due to genetic effects on BMI and aging. Overall, these results support a role of BMI in aging, potentially in part due to the effects of insulin resistance.

## INTRODUCTION

Obesity is a global public health which continues to increase in prevalence worldwide^1^. It contributes to numerous adverse health outcomes including cardiovascular disease^2^, diabetes^3^, and cancers^4,5^, all of which are considered diseases of aging. It is possible to estimate an individual’s age from DNA methylation (DNAm) at selected genomic sites using algorithms known as epigenetic “clocks”. These clocks include the original Horvath clock^6^, PhenoAge^7^, and HannumAge^8^, as well as the newer GrimAge which is more predictive of mortality than previous epigenetic clocks^9^ and thus may be a more appropriate measure of biological age. The difference between the predicted epigenetic age and chronological age is referred to as “age acceleration”, a phenomenon that occurs in the context of many diseases such as cancers.^6,10,11^ Additionally, associations of age acceleration with high BMI and obesity have been reported in some,^12-15^ but not all, studies^16,17^ when epigenetic aging is measured in blood. Two recent studies have identified associations between age acceleration based on GrimAge with BMI as well as associated clinical measures, such as triglycerides.^18,19^ Therefore, excess body mass may play a role in the heightened risk of conditions including cancers and cardiovascular disease experienced in obesity.

However, genotype is another important influence on both body composition and the epigenome,^20,21^ which introduces the possibility of genetic confounding in the assessment of the association between BMI and epigenetic aging. A monozygotic (MZ) co-twin control study design controls for genotype as well as sex and a variety of environmental exposures and experiences shared by MZ twin siblings. Here, we assessed the cross-sectional association of BMI with epigenetic age acceleration determined using the GrimAge clock in twins participating in the Finnish Twin Cohort, and two independent Finnish cohorts. To assess genetic confounding, we compared within-pair analyses including dizygotic (DZ) twin pairs and those limited to MZ twin pairs.

## MATERIALS AND METHODS

### Participants and study design

Study participants were MZ and DZ twin pairs participating in the Finnish Twin Cohort, comprised of three longitudinal cohorts. The Older Twin Cohort consists of same-sex twin pairs born before 1958, while FinnTwin12 and FinnTwin16 are longitudinal studies of five consecutive birth cohorts of Finnish twins born between 1975-1979 and 1983-1987,^22,23^ respectively; the two latter studies include opposite-sex twin pairs. Participants completed multiple surveys on behavioral and lifestyle traits as well as anthropometric measurements. Participants were selected for the current analysis if they had available data for blood DNA methylation, sex, zygosity, and concurrent height, weight, and age values, resulting in 1447 participants. MZ, same-sex DZ, and opposite-sex DZ twin pairs were included. A subset of MZ twin pairs participated in the TwinFat sub-cohort^24,25^ (n = 90 pairs), in which more detailed information on body composition and markers of cardiometabolic health including fat percentage, subcutaneous fat, intra-abdominal fat, liver fat percentage, and fasting total, LDL, and HDL cholesterol, triglycerides, C-reactive protein (CRP), leptin, adiponectin, glucose, and insulin as well as the homeostatic model assessment of insulin resistance (HOMA-IR) was available. All participants gave informed consent for their participation, and the study procedures were approved by the ethics committees of Helsinki University Central Hospital (113/E3/2001, 249/E5/2001, 346/E0/05, 270/13/03/01/2008 and 154/13/03/00/2011).

### Collection of biospecimens and DNA methylation measurement

Twins provided blood samples as part of targeted studies.^22,23^ As described previously, DNA was extracted from whole blood using the QIAamp DNA Mini kit (QIAGEN Nordic, Sollentuna, Sweden), and bisulfite conversion was performed with the EZ-96 DNA Methylation-Gold Kit (Zymo Research, Irvine, CA, USA) as per manufacturer instructions. We used the Illumina Infinium HumanMethylation450 BeadChip to measure DNA methylation at more than 480,000 CpGs site throughout the genome^26^. Samples from twin pairs were converted on the same plate in order to reduce batch effects due to technical variation.

### Quality control and preprocessing of DNA methylation data

Sample processing was completed in R version 3.6.0. Samples with poor quality were identified using the R package *MethylAid* with default thresholds^27^; those with a median methylated and unmethylated log_2_ intensity smaller than 10.5, an average log_2_ intensity of green and red channels’ expected signals of non-polymorphic controls smaller than 11.75, an average log_2_ intensity of converted bisulfite type I controls in green and red channels smaller than 12.75, an average log_2_ intensity of high and low hybridization controls (green channel) smaller than 13.25, or with less than 95% of probes with a detection *p*-value < 0.05 were excluded. Next, we normalized the DNA methylation data using *minfi*^*28*^. Removing bad quality samples resulted in a sample size of 1424. We performed functional normalization including the first two principal components of the control probes with noob background correction in order to reduce technical variation in the data^29^. We removed probes with a detection *p*-value > 0.01, an intensity value of exactly 0, or a bead count < 3 in more than 5% of samples. Beta-mixture quantile normalization was used to adjust beta values for differences due to probe type^30^ using the R package *wateRmelon*^*31*^. We additionally removed probes on sex chromosomes, and those identified as unreliable such as due to cross-reactivity^32^.

### Epigenetic age calculation

In this study, we used the newly developed epigenetic clock “GrimAge” developed by the Horvath group^9^, which is a DNA methylation-based biomarker of mortality. The GrimAge value is calculated in a two-step process, first estimating 7-plasma proteins including adrenomedullin, beta-2 microglobulin, cystatin, growth differentiation factor 15, leptin, plasminogen activation inhibitor 1, and tissue inhibitor metalloproteinase 1, as well as pack-years, from DNA methylation data, then using these estimates in combination with age, sex, and estimated smoking pack-years in a model developed from Cox proportional hazards regression. The resulting GrimAge estimate is scaled to be in units of years, with a higher GrimAge value corresponding to higher hazard of death.

First, we subset the data to only include probes used in estimating GrimAge. Any required probes with missing beta values were replaced with a beta value from the “gold database” provided by the Horvath group. Next, we added participant age and sex to the dataset, which are used in the estimation process. A python script provided by the Horvath group was used to estimate GrimAge, smoking pack-years, and the seven plasma proteins predicted in the first stage of estimation. Finally, age acceleration was calculated for each participant by regressing GrimAge on chronological age and taking the raw residual. Participants with a negative value of age acceleration have a lower epigenetic age than expected based on their chronological age, whereas those with positive age acceleration values have a higher epigenetic age.

### Statistical analysis

We used two approaches to assess the relationship between BMI and age acceleration, (1) treating each twin as an observation, and (2) treating twin pairs as observation. For both approaches, we used linear mixed effects models implemented in R version 3.2.2 and the R package *lme4*^*33*^ (version 1.1-11).

In the first case we accounted for the dependency within twin pairs by including a random intercept for family id, and additionally included random intercepts for twin cohort and zygosity. The dependent variable was age acceleration, while the independent variable was either BMI as a continuous measure, BMI as a categorical measure, or one of 14 clinical obesity-related measures as continuous measures (fat percentage, subcutaneous fat, intra-abdominal fat, liver fat percentage, and fasting total, LDL, HDL cholesterol, triglycerides, CRP, leptin, adiponectin, glucose and insulin as well as HOMA-IR). Clinical variables with non-normal distributions were transformed using the natural logarithm. Three adjusted versions of the models were performed, (1) adjusting for age, sex, and predicted smoking pack-years, (2) additionally adjusting for predicted proportions of CD8 T cells, CD4 T cells, natural killer cells, and neutrophils^34^, and (3) in the case of the clinical models, additionally adjusting for BMI. For the within-pair analyses, intrapair differences in epigenetic age, BMI, and covariates (predicted smoking pack years and cell type proportions) were calculated. Linear mixed effects models were performed with the dependent and independent variables being difference in epigenetic age and difference in BMI within a twin pair, respectively, adjusted for age, sex, and differences in predicted smoking pack-years and in fully adjusted models the same cell type proportions as in previous models, with random intercepts for zygosity and twin cohort. All *p*-values resulting from linear mixed effects models were calculated using the likelihood ratio test comparing the full model with the nested model. We considered *p*-values < 0.05 to be statistically significant.

Additional analyses were performed in R version 3.6.0. In order to validate our results, we next analyzed two independent cohorts, the Dietary, Lifestyle, and Genetic Determinants of Obesity and Metabolic Syndrome (DILGOM, n = 305) study and the Young Finns Study^35^ (YFS, n = 1581). DNA methylation was measured in blood using the Illumina 450k array in DILGOM, while both the 450k and EPIC array was used in the YFS. The quality control procedure described above was used for DILGOM and YFS data. The DILGOM data was preprocessed using the same steps as in FTC data; in order to combine the 450k and EPIC data into a single dataset for the YFS the array probes were limited to those present on both the 450k and EPIC arrays. GrimAge and age acceleration were calculated as described above. We used linear regression to assess the association between BMI or clinical measures and age acceleration, adjusting for age, sex, and predicted smoking pack-years. We additionally adjusted for predicted proportions of blood cell types as in the FTC analyses.

We performed a meta-analysis for BMI and the clinical variables overlapping between studies using the individual-level results from the FTC, YFS, and DILGOM to obtain the best estimates for the effects of each variable on insulin resistance. Random-effect meta-analyses were performed using the R libraries *meta* and *metafor*. The empirical Bayes method was used for estimating the between-study variance.

## RESULTS

### Participant characteristics

Participant characteristics for all participants are presented in **Table 1**. There were a total of 1424 twin individuals from the FTC included in this study, with 790 MZ twins, 445 same-sex DZ twins, and 189 opposite-sex DZ twins; additionally, there were non-twin participants from the YFS (n = 1591) and DILGOM (n = 304). In the FTC, age ranged from 21-73 years old, with an average age of 34.5 years, while the age ranged from 34-49 years in the YFS and 25-74 years in DILGOM. A majority of participants were female in all three studies (57.7%, 55.6%, and 52.9%). The average BMI was lowest in the FTC at 24.7 kg/m^2^ compared to 26.6 in both the YFS and DILGOM.

**Table 1.**
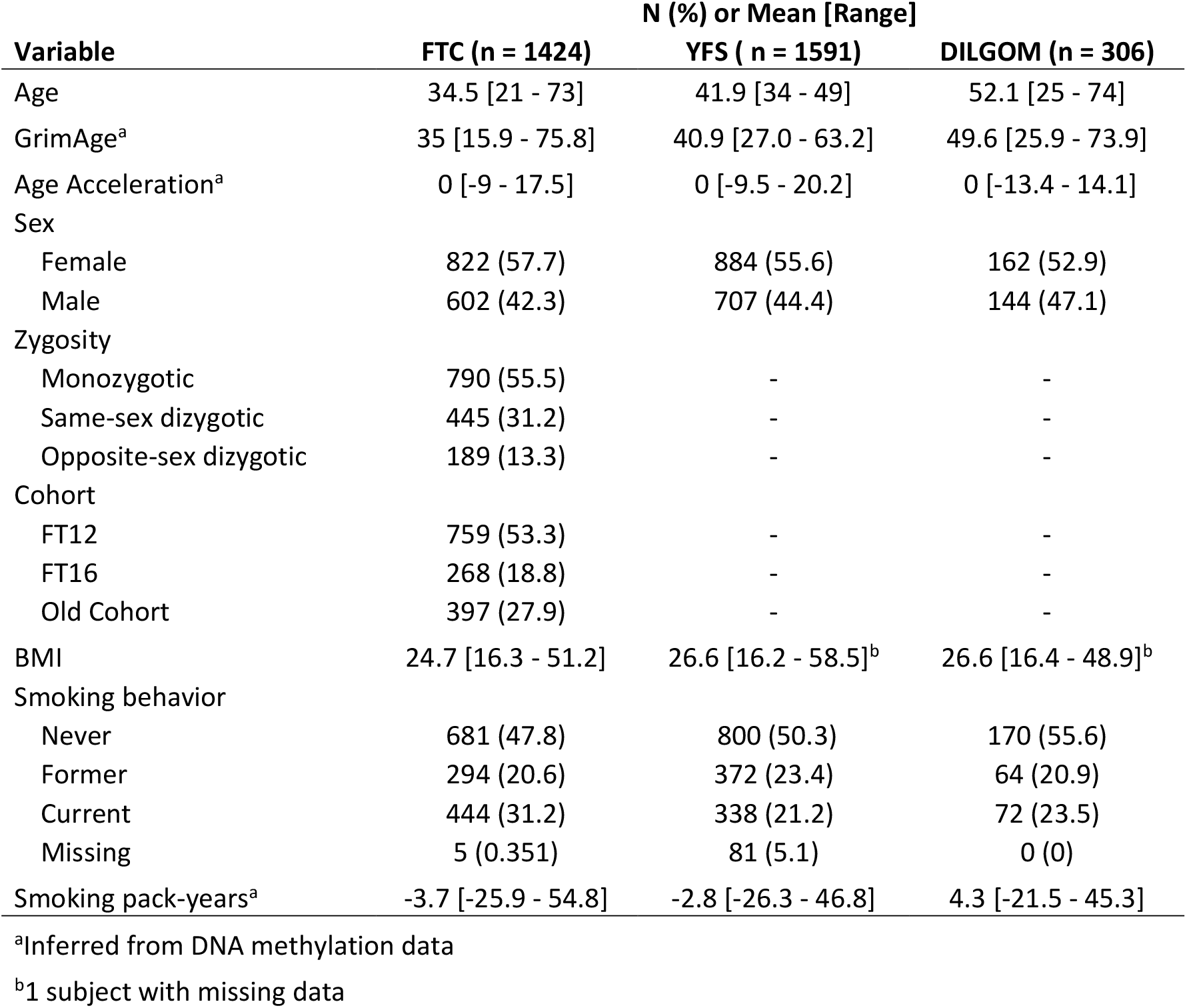
Participant Characteristics.

| Variable | N (%) or Mean [Range] |  |  |
| --- | --- | --- | --- |
|  | FTC (n = 1424) | YFS ( n = 1591) | DILGOM (n = 306) |
| Age | 34.5 [21 - 73] | 41.9 [34 - 49] | 52.1 [25 - 74] |
| GrimAge<sup>a</sup> | 35 [15.9 - 75.8] | 40.9 [27.0 - 63.2] | 49.6 [25.9 - 73.9] |
| Age Acceleration<sup>a</sup> | 0 [-9 - 17.5] | 0 [-9.5 - 20.2] | 0 [-13.4 - 14.1] |
| Sex |  |  |  |
| Female | 822 (57.7) | 884 (55.6) | 162 (52.9) |
| Male | 602 (42.3) | 707 (44.4) | 144 (47.1) |
| Zygoty |  |  |  |
| Monozygotic | 790 (55.5) | - | - |
| Same-sex dizygotic | 445 (31.2) | - | - |
| Opposite-sex dizygotic | 189 (13.3) | - | - |
| Cohort |  |  |  |
| FT12 | 759 (53.3) | - | - |
| FT16 | 268 (18.8) | - | - |
| Old Cohort | 397 (27.9) | - | - |
| BMI | 24.7 [16.3 - 51.2] | 26.6 [16.2 - 58.5]<sup>b</sup> | 26.6 [16.4 - 48.9]<sup>b</sup> |
| Smoking behavior |  |  |  |
| Never | 681 (47.8) | 800 (50.3) | 170 (55.6) |
| Former | 294 (20.6) | 372 (23.4) | 64 (20.9) |
| Current | 444 (31.2) | 338 (21.2) | 72 (23.5) |
| Missing | 5 (0.351) | 81 (5.1) | 0 (0) |
| Smoking pack-years<sup>a</sup> | -3.7 [-25.9 - 54.8] | -2.8 [-26.3 - 46.8] | 4.3 [-21.5 - 45.3] |

<sup>a</sup>Inferred from DNA methylation data
<sup>b</sup>1 subject with missing data

### Individual analysis

First, we assessed the relation of BMI as a continuous measure with age acceleration with twins as individuals. Each 1-unit increase in BMI corresponded to an increase in age acceleration of 1.02 months (likelihood ratio *p*-value = 6.1 × 10^−12^, **Figure 1a**). After adjusting for cell type proportions, the effect of BMI on age acceleration was slightly attenuated, with each 1-unit increase in BMI corresponding to 0.91 months (likelihood ratio *p*-value = 9.0 × 10^−11^). BMI was positively associated with age acceleration in both validation populations, with a one-unit increase in BMI associated with an increase in age acceleration of 0.67 months in DILGOM (*p*-value = 0.0050) and 1.08 months in the YFS (*p*-value = 2.6 × 10-^27^). Performing a meta-analysis of these estimates revealed no evidence of heterogeneity between studies (*p*-value = 0.23), and an overall estimate for the effect of BMI on age acceleration of 0.08 years or 0.96 months per each 1-unit BMI increase (**Figure 1b**).

**Figure 1.**
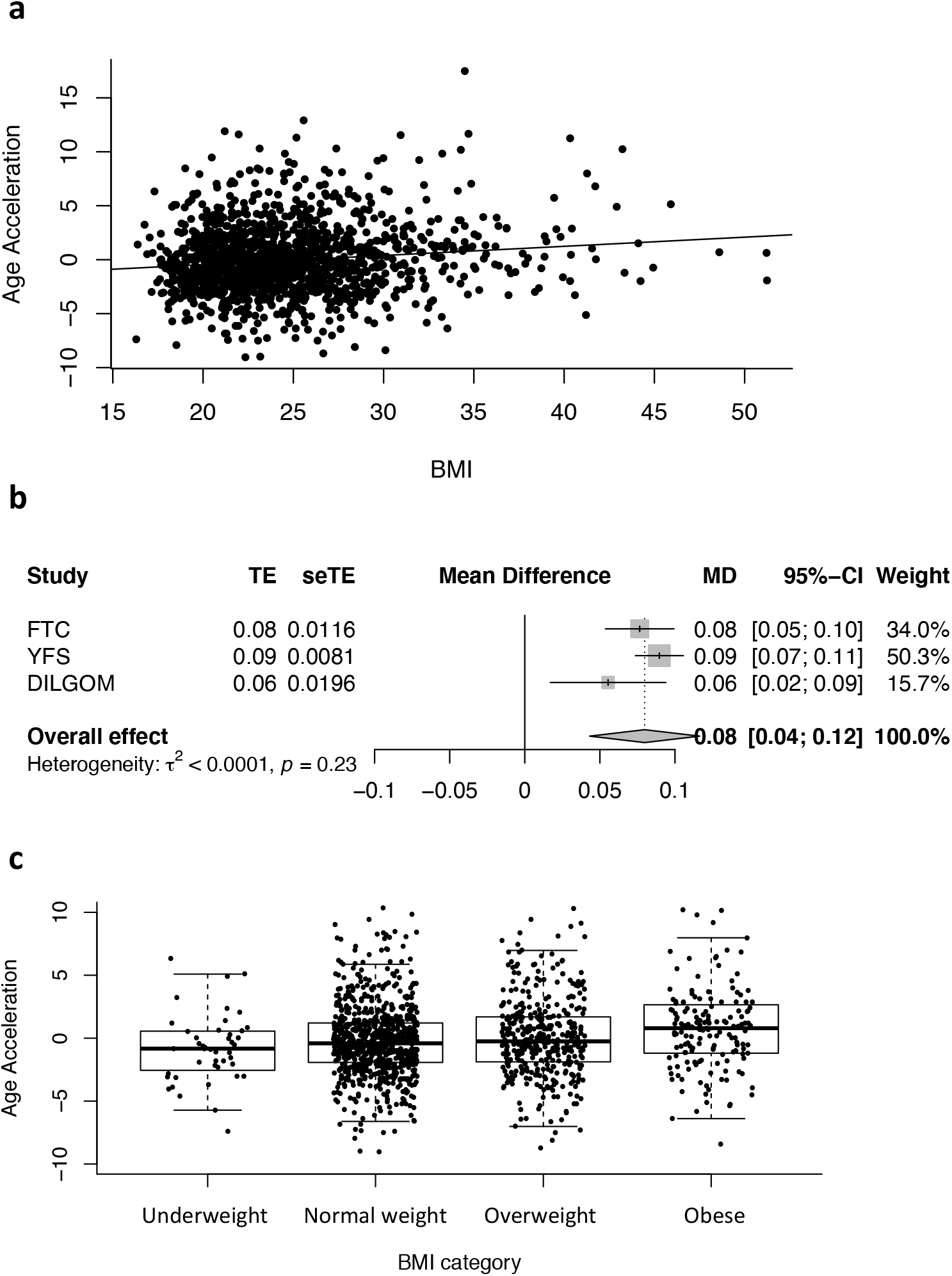
BMI associates with age acceleration. (a) Scatterplot with best fit line showing the association of age acceleration with BMI in the FTC. (b) Forest plot showing the estimates for the association of BMI with age acceleration in each study, and the overall effect from a meta-analysis. (c) Boxplot showing differences in age acceleration by BMI category.

We observed a linear association of BMI categories with age acceleration; compared to individuals classified as underweight, the age acceleration of normal weight participants was 6.6 months higher, that of overweight participants was 10.9 months higher, and that of obese participants was 1.6 years higher (likelihood ratio *p*-value = 8.9 × 10^−9^, **Figure 1c**). This association was attenuated after adjusting for blood cell type proportions, but the association remained consistent in direction and significance.

### Differences in epigenetic aging within twin pairs

Next, we calculated the differences in BMI, and epigenetic age between each twin in a pair, subtracting the lighter twin from the heavier twin. Each 1-unit increase in BMI in the heavier twin was associated with an increase in epigenetic age of 1.6 months compared to their lighter co-twin (likelihood ratio *p*-value = 4.7 × 10^−12^; **Figure 2a**). There was no heterogeneity by zygosity, with the estimate for the random effect at 0. We repeated the same analysis including only MZ twin pairs in order to fully control for the effect of genetics, which showed that each 1-unit difference in BMI within the twin pairs was associated with an increase in epigenetic age of 1.1 months (likelihood ratio *p*-value = 0.00010; **Figure 2b**).

**Figure 2.**
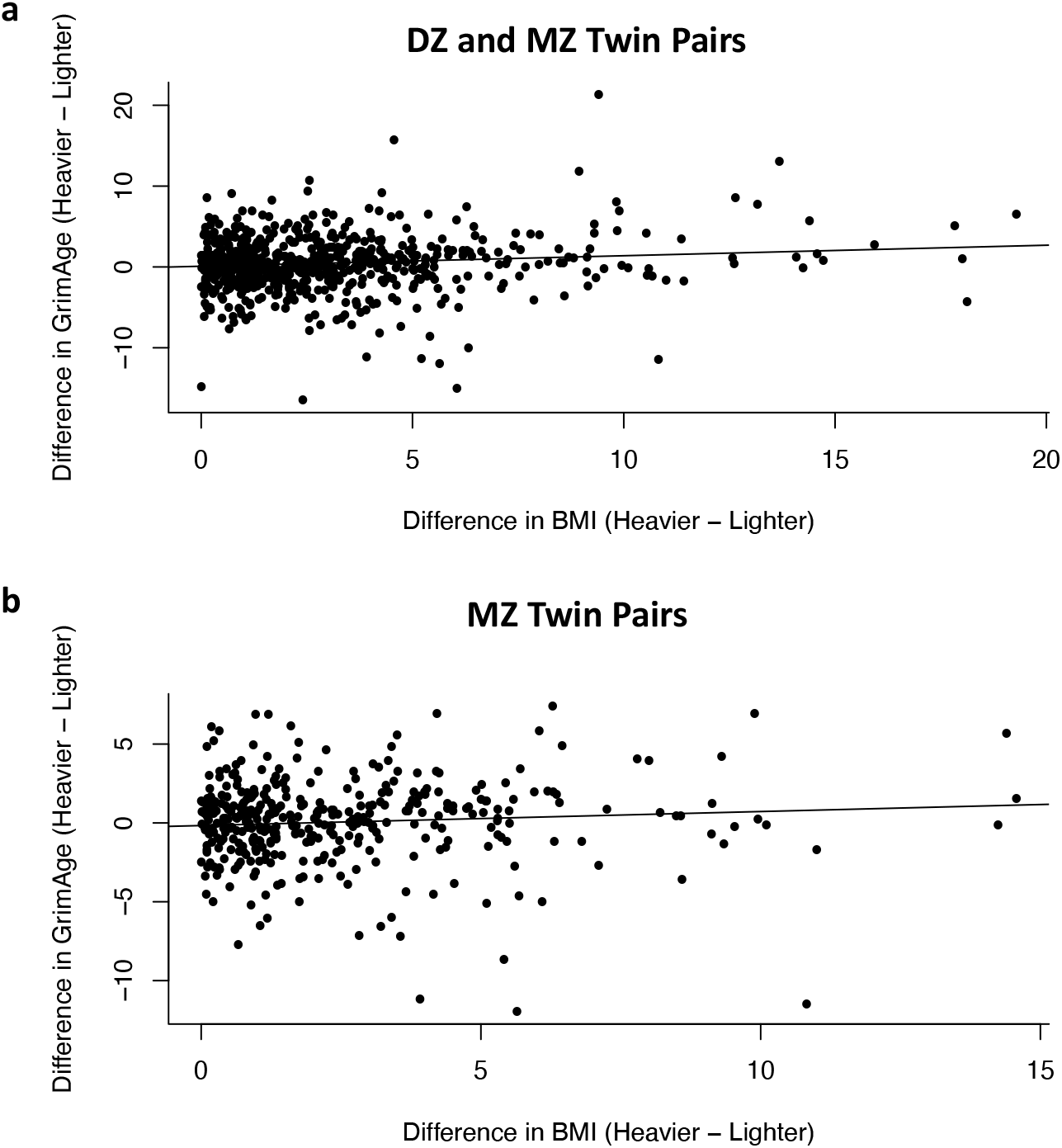
Difference in BMI is related with the difference in GrimAge within twin pairs. (a) Scatterplot showing the association of the difference in BMI within twin pairs in relation with the difference in GrimAge, including both DZ and MZ twin-pairs. One twin pair was excluded due to an extreme difference in BMI of greater than 30. (b) Scatterplot showing the association of the difference in BMI within twin pairs in relation with the difference in GrimAge, including only MZ twin-pairs.

### BMI discordance within monozygotic twin pairs

The average BMI of the lighter twins in BMI-discordant pairs was 24.9 kg/m^2^, versus 30.3 kg/m^2^ for the heavier twins, and the average age was 41.9 years. The heavier twins were less likely to smoke, with 24.8% of heavier twins current smokers, versus 31.6% of lighter twins. Heavier co-twins had higher age acceleration by 5.2 months compared to their leaner co-twin (likelihood ratio *p*-value = 0.0066; **Figure S1**). The average difference in BMI between a discordant pair was around 5 units, resulting in an effect size per BMI-unit of 1.04 months.

### Clinical measures and age acceleration

A subset of 90 monozygotic twins belonging to BMI-discordant twin pairs were evaluated clinically for obesity-related measures. In the meta-analysis, both log(HOMA-IR index) and log(fasting insulin) were significantly associated with age acceleration both before and after adjusting for BMI (**Tables 2-4, Figure 3**). There was no evidence of heterogeneity between studies for HOMA-IR or fasting insulin (**Figure 3**). Additionally, log(CRP) was positively associated with age acceleration in YFS participants, both before and after adjusting for BMI (**Table 4**), but was only significantly associated with age acceleration in FTC participants in the model not adjusting for cell types or BMI (**Table 2**). This association was marginally significant in the meta-analysis for the models additionally adjusting for BMI (Coefficient = 0.23 years, *p*-value = 0.053). Other clinical measures were not significantly associated with age acceleration in meta-analyses. HDL cholesterol was consistently negatively associated with age acceleration, but this was only significant in the YFS, and there was a significant amount of heterogeneity between studies (*p*-value = 0.036). Measures of body fat were only available in FTC participants; each dm^3^ increase in subcutaneous fat was positively associated with an acceleration in age of 2.79 months (*p*-value = 0.0031), each log(intra-abdominal fat dm^3^) was associated with an increased epigenetic age of 5.63 months (*p*-value = 0.028), and a unit increase in log(liver fat %) corresponded to increased age acceleration of 3.29 months (*p*-value = 0.047).

**Table 2.**
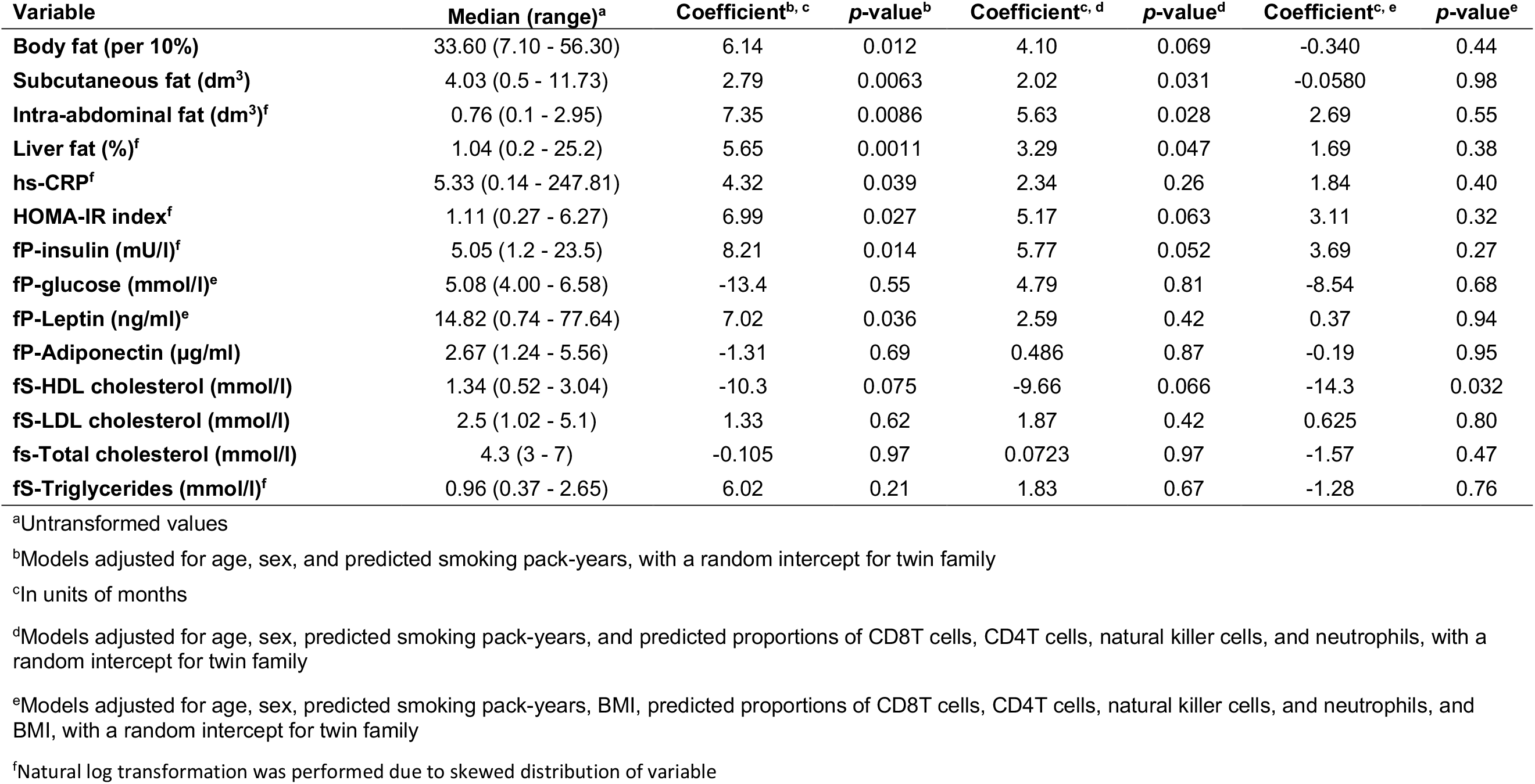
Obesity-related clinical measures associate with age acceleration (n = 90).

**Table 3.** BMI and obesity-related clinical measures in relation to age acceleration in DILGOM participants (n = 305).

| Variable | Median (range)<sup>a</sup> | Coefficient<sup>b, c</sup> | p-value<sup>b</sup> | Coefficient<sup>c, d</sup> | p-value<sup>d</sup> | Coefficient<sup>c, e</sup> | p-value<sup>e</sup> |
| --- | --- | --- | --- | --- | --- | --- | --- |
| **BMI** | 25.8 (16.4 - 48.9) | 0.59 | 0.019 | 0.67 | 0.0050 | - | - |
| **fP-glucose (mmol/l)<sup>f</sup>** | 5.8 (4.4 - 15.0) | -13.8 | 0.14 | -6.78 | 0.44 | -11.5 | 0.19 |
| **fP-insulin (mU/l)<sup>f</sup>** | 5.3 (1.5 - 94.5) | 5.26 | 0.012 | 5.52 | 0.0044 | 3.61 | 0.12 |
| **HOMA-IR Index<sup>f</sup>** | 1.33 (0.34 - 23.53) | 3.65 | 0.051 | 4.16 | 0.017 | 2.17 | 0.29 |
| **fS-HDL cholesterol (mmol/l)** | 1.42 (0.51 - 2.82) | -3.26 | 0.32 | -3.23 | 0.30 | -1.15 | 0.72 |
| **fS-Triglycerides (mmol/l)<sup>f</sup>** | 1.02 (0.37 - 5.14) | 2.3 | 0.39 | 4.6 | 0.068 | 2.59 | 0.33 |

<sup>a</sup>Untransformed values
<sup>b</sup>Models adjusted for age, sex, and predicted smoking pack-years
<sup>c</sup>In units of months
<sup>d</sup>Models adjusted for age, sex, predicted smoking pack-years, and predicted proportions of CD8T cells, CD4T cells, natural killer cells, and neutrophils
<sup>e</sup>Models adjusted for age, sex, predicted smoking pack-years, predicted proportions of CD8T cells, CD4T cells, natural killer cells, and neutrophils, and BMI
<sup>f</sup>Natural log transformation was performed due to skewed distribution of variable

**Table 4.** Association of BMI and obesity-related clinical measures with age acceleration in YFS (n = 1591).

| Variable | Median (range)<sup>a</sup> | Coefficient<sup>b, c</sup> | p-value<sup>b</sup> | Coefficient<sup>c, d</sup> | p-value<sup>d</sup> | Coefficient<sup>c, e</sup> | p-value<sup>e</sup> |
| --- | --- | --- | --- | --- | --- | --- | --- |
| **BMI** | 25.8 (16.2 - 58.5) | 1.14 | 6.6E-27 | 1.08 | 2.6E-27 | - | - |
| **HbA1C<sup>f</sup>** | 36 (22 - 102) | 15.2 | 0.0018 | 18.7 | 3.7E-05 | 4.89 | 0.29 |
| **fP-insulin (mU/l)<sup>f, g</sup>** | 7.41 (0.06 - 95.7) | 6.30 | 2.0E-21 | 5.90 | 1.6E-21 | 3.14 | 1.7E-05 |
| **fP-glucose (mmol/l)<sup>f, h</sup>** | 5.25 (3.14 - 12.65) | 34.6 | 1.7E-12 | 33.2 | 3.7E-13 | 19.9 | 2.0E-05 |
| **fs-HDL cholesterol (mmol/l)** | 1.29 (0.52 - 2.64) | -9.69 | 1.6E-08 | -8.99 | 2.6E-08 | -3.77 | 0.024 |
| **fs-LDL cholesterol (mmol/l)** | 3.19 (1.06 - 7.05) | -0.0628 | 0.93 | -0.00515 | 0.99 | -0.552 | 0.37 |
| **fs-Total cholesterol (mmol/l)** | 5.1 (2.8 - 10.2) | 0.450 | 0.44 | 0.802 | 0.14 | 0.222 | 0.68 |
| **fs-Triglycerides (mmol/l)<sup>f</sup>** | 1.05 (0.34 - 6.05) | 7.95 | 1.3E-13 | 8.92 | 5.4E-19 | 5.52 | 2.0E-07 |
| **hs-CRP<sup>f</sup>** | 0.79 (0.05 - 29.08) | 5.16 | 9.9E-32 | 4.51 | 1.7E-27 | 3.02 | 1.1E-10 |
| **HOMA-IR index<sup>f</sup>** | 1.73 (0.02 - 21.0) | 5.82 | 4.8E-22 | 5.49 | 1.9E-22 | 3.03 | 4.7E-06 |

<sup>a</sup>Untransformed values
<sup>b</sup>Models adjusted for age, sex, and predicted smoking pack-years
<sup>c</sup>In units of months
<sup>d</sup>Models adjusted for age, sex, predicted smoking pack-years, and predicted proportions of CD8T cells, CD4T cells, natural killer cells, and neutrophils
<sup>e</sup>Models adjusted for age, sex, predicted smoking pack-years, predicted proportions of CD8T cells, CD4T cells, natural killer cells, and neutrophils, and BMI
<sup>f</sup>Natural log transformation was performed due to skewed distribution of variable
<sup>g</sup>1 sample excluded for extreme logtransformed insulin value (mU/l) of over 8
<sup>h</sup>2 samples excluded for extreme log-transformed glucose (mmol/l) value of over 2.75

**Figure 3.**
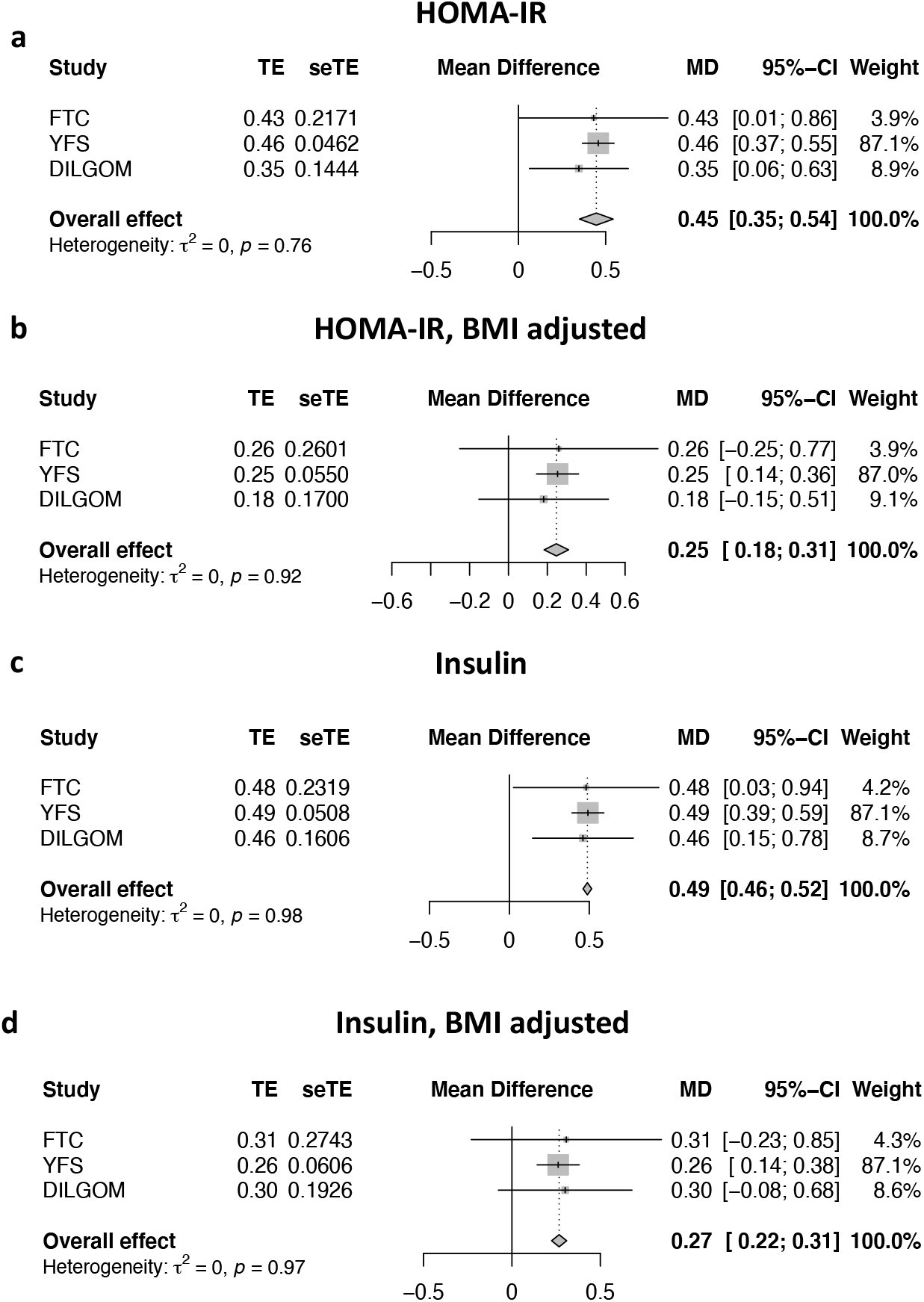
Obesity-related clinical measures associate with age acceleration. Forest plots showing positive associations between (a) HOMA-IR index without BMI adjustment, (b) HOMA-IR index with BMI adjustment, (c) fasting insulin without BMI adjustment, and (d) fasting insulin with BMI adjustment in meta-analyses including all three cohorts. Estimates are in units of years.

## DISCUSSION

In this study we investigated the association of BMI with epigenetic aging using a new epigenetic clock, GrimAge, using Finnish MZ and DZ twin pairs. We found a positive association between BMI and age acceleration as inferred from DNA methylation in blood. Importantly, the observation of epigenetic age acceleration between heavy and light twins within BMI-discordant MZ twin pairs shows that this association is not due to confounding by shared genetic and environmental effects on DNA methylation at aging-associated CpGs. Additionally, we found a linear relationship between BMI and age acceleration, with underweight individuals displaying the lowest amount of age acceleration, and an incremental increase in the amount of age acceleration through each subsequent BMI category. This is noteworthy given that most studies of BMI suggest that underweight individuals are at higher risk of disease,^36-38^ however we find no evidence of that here. Finally, we found that the most strongly associated obesity-related clinical features were those related to glucose metabolism and insulin resistance, as shown by differences in age acceleration in relation to the HOMA-IR index and fasting insulin, even after additionally adjusting for BMI.

The association of BMI with accelerated epigenetic aging was also observed in two independent Finnish cohorts of unrelated individuals, DILGOM and the YFS. However, the effect estimates for the relation of BMI with age acceleration were slightly different in the FTC compared to DILGOM and YFS. The analysis including all twins showed an increase in age acceleration of 1.7 months per unit BMI increase among all individual participants, while within BMI discordant MZ twin pairs whose co-twins differed in BMI on average by 5 BMI units showed an increase of 5.2 months in age acceleration, or around 1 month per unit BMI difference. However, DILGOM participants displayed an age acceleration increase of around 0.7 months and YFS participants around 1 month per unit BMI increase. Nonetheless, our meta-analysis provides no evidence of heterogeneity between studies for the effect estimate of BMI on age acceleration. Interestingly, we observed associations of HOMA-IR index with age acceleration in all three cohorts of which the magnitude was comparable in all three studies, with the meta-analyses not indicating any heterogeneity. The lack of evidence of heterogeneity in the associations of BMI and measures of glucose metabolism with age acceleration may indicate that genetics do not strongly confound these associations. The association is thus robust to differences in the ascertainment of the samples, and can be considered to be a true population effect.

Based on the results we obtained, insulin resistance may be responsible in part for the effect of obesity on epigenetic aging, since adjusting for BMI results in a reduction in the effect estimate of HOMA-IR on age acceleration. Obesity and aging both play a role in insulin resistance and type 2 diabetes^39-41^. Obesity is known to promote inflammation^42^, which in turn is involved in the onset of lipid-induced insulin resistance^41^. Interestingly, metformin, a drug used to lower blood glucose levels, is being tested as an intervention to protect against aging and age-related diseases^43^. Additionally, we also observed a trend of increased age acceleration with rising CRP, an inflammatory marker, however this association disappeared after adjusting analyses for the predicted proportion of immune-cell types.

These findings are consistent with other studies using GrimAge to determine age acceleration,^18,19^ however other epigenetic clocks seem to be able to detect this effect better in other tissues such as liver^17^ and visceral adipose tissue^16^ than blood. This could indicate that the GrimAge epigenetic clock is better suited for usage for assessment of age acceleration related to cardiometabolic phenotypes, which is possible given the unique process used to develop the GrimAge clock of estimating blood proteins. This includes leptin, for example, which is known to be higher in obesity^44^. Overall, our results support the well-established association of BMI with aging as well as a strong role of insulin resistance.

Further, nutrient-sensing pathways may play a role in the relation of obesity and aging. For example, genes belonging to nutrient-sensing pathways including insulin/insulin-like growth factor (IGF) pathway, mechanistic target of rapamycin (mTOR), adenosine monophosphate-activated protein kinase (AMPK), and sirtuin deacetylases appear to regulate lifespan in mice^45^. In fact, mutations in these genes including IGF1 and IGFR associate with increased longevity in humans,^46^ and a low energy state activates AMPK as well as sirtuins^47^. This information points to a link between nutrient-sensing pathways, weight gain, and aging, since weight gain is caused in part by excessive energy intake.

Our study has several strengths which contribute to its significance. First, the usage of MZ twin pairs discordant for obesity allows us to be certain that the associations we identified are not entirely due to confounding by genetic predisposition to both obesity and accelerated aging. Additionally, we identified associations with obesity-related clinical measures that are in line with the results obtained for BMI, with a detrimental effect of insulin resistance and a beneficial effect of HDL on aging. Further, we performed meta-analyses for the associations of BMI and obesity-related clinical measures with age acceleration, and demonstrated consistent associations for BMI, HOMA-IR, and fasting insulin with increased age acceleration in all three studies. However, our study is limited by the small number of MZ twin pairs discordant for BMI, which is due to the rarity of this occurring. Additionally, our study populations consisted of exclusively Finnish participants, which may somewhat limit the generalizability of our findings to other populations although this is unlikely given that the same associations have previously been observed in other populations.

## CONCLUSION

In conclusion, we identified significant associations of BMI, HOMA-IR, a measure of insulin resistance, and fasting insulin with epigenetic age acceleration calculated using the GrimAge epigenetic clock. These associations were not due the effects of genetics on BMI and aging, and measures of insulin resistance were also associated independently from BMI with age acceleration. Overall, these results suggest that BMI plays a role in aging, along with and perhaps in part due to insulin resistance. More research needs to be done to determine if weight loss can reverse BMI-associated epigenetic aging.

## Data Availability

The data are not publicly available. Please contact for inquiries about the data.

## Author contributions

MO and SK conceptualized the aims of the study, SL and SK performed statistical analyses and created figures for the FTC data, SL performed statistical analyses and created figures in the validation populations, SL and SK wrote the paper, and all authors aided in the interpretation of results, offered critical feedback, and reviewed and edited the paper.

---

**Figure S1.**
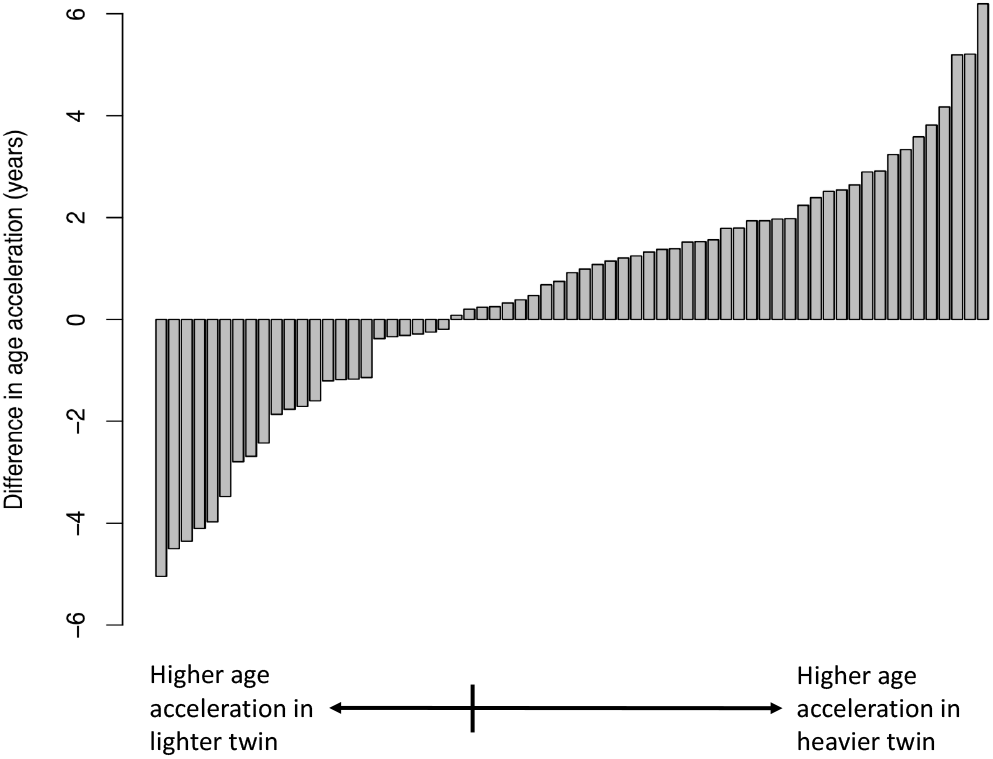
Heavier twins in a BMI-discordant pair more often have higher age acceleration compared to their leaner co-twin. Waterfall plot showing the difference in age acceleration in years within MZ twin pairs discordant for obesity, subtracting the value for the leaner twin from that of the heavier twin.

## REFERENCES

1 Peralta, M., Ramos, M., Lipert, A., Martins, J. & Marques, A. Prevalence and trends of overweight and obesity in older adults from 10 European countries from 2005 to 2013. Scandinavian journal of public health 46, 522–529 (2018).

2 Collaboration, E. R. F. Separate and combined associations of body-mass index and abdominal adiposity with cardiovascular disease: collaborative analysis of 58 prospective studies. The Lancet 377, 1085–1095 (2011).

3 Narayan, K. V., Boyle, J. P., Thompson, T. J., Gregg, E. W. & Williamson, D. F. Effect of BMI on lifetime risk for diabetes in the US. Diabetes care 30, 1562–1566 (2007).

4 Wolk, A. et al. A prospective study of obesity and cancer risk (Sweden). Cancer causes & control 12, 13–21 (2001).

5 Bardou, M., Barkun, A. N. & Martel, M. Obesity and colorectal cancer. Gut 62, 933–947 (2013).

6 Horvath, S. DNA methylation age of human tissues and cell types. Genome biology 14, 3156 (2013).

7 Levine, M. E. et al. An epigenetic biomarker of aging for lifespan and healthspan. Aging (Albany NY) 10, 573–591, doi:10.18632/aging.101414 (2018).

8 Hannum, G. et al. Genome-wide methylation profiles reveal quantitative views of human aging rates. Mol Cell 49, 359–367, doi:10.1016/j.molcel.2012.10.016 (2013).

9 Lu, A. T. et al. DNA methylation GrimAge strongly predicts lifespan and healthspan. Aging (Albany NY) 11, 303 (2019).

10 Yang, Z. et al. Correlation of an epigenetic mitotic clock with cancer risk. Genome biology 17, 1–18 (2016).

11 Perna, L. et al. Epigenetic age acceleration predicts cancer, cardiovascular, and all-cause mortality in a German case cohort. Clinical epigenetics 8, 64 (2016).

12 Huang, R.-C. et al. Epigenetic age acceleration in adolescence associates with BMI, inflammation, and risk score for middle age cardiovascular disease. The Journal of Clinical Endocrinology & Metabolism 104, 3012–3024 (2019).

13 Nevalainen, T. et al. Obesity accelerates epigenetic aging in middle-aged but not in elderly individuals. Clinical epigenetics 9, 20 (2017).

14 Quach, A. et al. Epigenetic clock analysis of diet, exercise, education, and lifestyle factors. Aging (Albany NY) 9, 419 (2017).

15 Ryan, J., Wrigglesworth, J., Loong, J., Fransquet, P. D. & Woods, R. L. A systematic review and meta-analysis of environmental, lifestyle, and health factors associated with DNA methylation age. The Journals of Gerontology: Series A 75, 481–494 (2020).

16 de Toro-Martín, J. et al. Body mass index is associated with epigenetic age acceleration in the visceral adipose tissue of subjects with severe obesity. Clinical epigenetics 11, 1–11 (2019).

17 Horvath, S. et al. Obesity accelerates epigenetic aging of human liver. Proceedings of the National Academy of Sciences 111, 15538–15543 (2014).

18 Arpón, A. et al. Interaction among sex, ageing and epigenetic processes concerning visceral fat, insulin resistance and dyslipidaemia. Frontiers in endocrinology 10, 496 (2019).

19 Zhao, W. et al. Education and lifestyle factors are associated with DNA methylation clocks in older African Americans. International journal of environmental research and public health 16, 3141 (2019).

20 Tekola-Ayele, F., Lee, A., Workalemahu, T. & Sánchez-Pozos, K. Shared genetic underpinnings of childhood obesity and adult cardiometabolic diseases. Human genomics 13, 17 (2019).

21 Smith, A. K. et al. Methylation quantitative trait loci (meQTLs) are consistently detected across ancestry, developmental stage, and tissue type. BMC genomics 15, 145 (2014).

22 Kaprio, J. The Finnish Twin Cohort Study: an update. Twin Res Hum Genet 16, 157–162, doi:10.1017/thg.2012.142 (2013).

23 Kaprio, J. et al. The Older Finnish Twin Cohort - 45 Years of Follow-up. Twin Res Hum Genet 22, 240–254, doi:10.1017/thg.2019.54 (2019).

24 Naukkarinen, J. et al. Characterising metabolically healthy obesity in weight-discordant monozygotic twins. Diabetologia 57, 167–176 (2014).

25 Heinonen, S. et al. Impaired Mitochondrial Biogenesis in Adipose Tissue in Acquired Obesity. Diabetes 64, 3135–3145, doi:10.2337/db14-1937 (2015).

26 Bibikova, M. et al. High density DNA methylation array with single CpG site resolution. Genomics 98, 288–295 (2011).

27 Van Iterson, M. et al. MethylAid: visual and interactive quality control of large Illumina 450k datasets. Bioinformatics 30, 3435–3437 (2014).

28 Aryee, M. J. et al. Minfi: a flexible and comprehensive Bioconductor package for the analysis of Infinium DNA methylation microarrays. Bioinformatics 30, 1363–1369 (2014).

29 Fortin, J.-P. et al. Functional normalization of 450k methylation array data improves replication in large cancer studies. Genome biology 15, 503 (2014).

30 Teschendorff, A. E. et al. A beta-mixture quantile normalization method for correcting probe design bias in Illumina Infinium 450 k DNA methylation data. Bioinformatics 29, 189–196 (2013).

31 Wong, C. C., Pidsley, R. & Schalkwyk, L. C. The wateRmelon Package. (2013).

32 Zhou, W., Laird, P. W. & Shen, H. Comprehensive characterization, annotation and innovative use of Infinium DNA methylation BeadChip probes. Nucleic acids research 45, e22–e22 (2017).

33 Bates, D., Maechler, M., Bolker, B., Walker, S. & Haubo Bojesen Christensen, R. (2015).

34 Salas, L. A. et al. An optimized library for reference-based deconvolution of whole-blood biospecimens assayed using the Illumina HumanMethylationEPIC BeadArray. Genome biology 19, 1–14 (2018).

35 Raitakari, O. T. et al. Cohort profile: the cardiovascular risk in Young Finns Study. Int J Epidemiol 37, 1220–1226, doi:10.1093/ije/dym225 (2008).

36 Aune, D. et al. BMI and all cause mortality: systematic review and non-linear dose-response meta-analysis of 230 cohort studies with 3.74 million deaths among 30.3 million participants. BMJ 353, i2156, doi:10.1136/bmj.i2156 (2016).

37 Sun, Y. Q. et al. Body mass index and all cause mortality in HUNT and UK Biobank studies: linear and non-linear mendelian randomisation analyses. BMJ 364, 1042, doi:10.1136/bmj.l1042 (2019).

38 Jayedi, A., Soltani, S., Zargar, M. S., Khan, T. A. & Shab-Bidar, S. Central fatness and risk of all cause mortality: systematic review and dose-response meta-analysis of 72 prospective cohort studies. BMJ 370, m3324, doi:10.1136/bmj.m3324 (2020).

39 Ehrhardt, N. et al. Adiposity-Independent Effects of Aging on Insulin Sensitivity and Clearance in Mice and Humans. Obesity 27, 434–443 (2019).

40 Salvestrini, V., Sell, C. & Lorenzini, A. Obesity may accelerate the aging process. Frontiers in endocrinology 10, 266 (2019).

41 Vorotnikov, A., Stafeev, I., Menshikov, M. Y., Shestakova, M. & Parfyonova, Y. V. Latent Inflammation and Defect in Adipocyte Renewal as a Mechanism of Obesity-Associated Insulin Resistance. Biochemistry (Moscow) 84, 1329–1345 (2019).

42 Karczewski, J. et al. Obesity and inflammation. European cytokine network 29, 83–94 (2018).

43 Barzilai, N., Crandall, J. P., Kritchevsky, S. B. & Espeland, M. A. Metformin as a tool to target aging. Cell metabolism 23, 1060–1065 (2016).

44 Izquierdo, A. G., Crujeiras, A. B., Casanueva, F. F. & Carreira M. C. Leptin, obesity, and leptin resistance: where are we 25 years later? Nutrients 11, 2704 (2019).

45 Singh, P. P., Demmitt, B. A., Nath, R. D. & Brunet, A. The genetics of aging: a vertebrate perspective. Cell 177, 200–220 (2019).

46 Suh, Y. et al. Functionally significant insulin-like growth factor I receptor mutations in centenarians. Proceedings of the National Academy of Sciences 105, 3438–3442 (2008).

47 López-Lluch, G. &Navas, P. Calorie restriction as an intervention in ageing. The Journal of physiology 594, 2043–2060 (2016).

